# Transmission event of SARS-CoV-2 Delta variant reveals multiple vaccine breakthrough infections

**DOI:** 10.1101/2021.06.28.21258780

**Authors:** Timothy Farinholt, Harsha Doddapaneni, Xiang Qin, Vipin Menon, Qingchang Meng, Ginger Metcalf, Hsu Chao, Marie-Claude Gingras, Paige Farinholt, Charu Agrawal, Donna M. Muzny, Pedro A. Piedra, Richard A. Gibbs, Joseph Petrosino

## Abstract

**Importance:** Vaccine breakthrough by an emergent SARS-CoV-2 variant poses a great risk to global public health.

**Objective:** To determine the SARS-CoV-2 variant responsible for 6 cases of vaccine breakthrough.

**Design:** Nasopharyngeal swabs from suspected vaccine breakthrough cases were tested for SARS-CoV-2 by qPCR for Wuhan-Hu1 and Alpha variant. Positive samples were then sequenced by Swift Normalase Amplicon Panels to determine the causal variant.

**Setting:** Transmission event occurred at events surrounding a wedding outside of Houston, TX. Two patients from India, likely transmitted the Delta variant to other guests.

**Participants:** Following a positive SARS-CoV-2 qPCR test at a third-party site, six fully vaccinated patients were investigated. Three males and three females ranged from 53 to 69 years old. One patient suffered from diabetes while three others were classified as overweight. No significant other comorbidities were identified. None of the patients had a history of failed vaccination.

**Key Points:** *Question:* Which SARS-CoV-2 variant is responsible for 6 cases of vaccine breakthrough, one interventional monoclonal antibody treatment, and one death?

*Findings:* Viral sequencing revealed 6 vaccinated patients were infected with the Delta SARS-CoV-2 variant. With no histories of vaccine breakthrough, this suggests Delta variant may possess immune evasion in patients that received the Pfizer BNT162b2, Moderna mRNA-1273, and Covaxin BBV152.

*Meaning:* Delta variant may pose the highest risk out of any currently circulating SARS-CoV-2 variants, with increased transmissibility over Alpha variant and possible vaccine breakthrough.

## Introduction

High numbers of global SARS-CoV-2 infections have led to the emergence of variants, notably Alpha variant (B.1.1.7 UK), Beta (B.1.351 S. Africa), Gamma (P.1 Brazil), Epsilon (B.1.429 California), Iota (B.1.526 New York) and now, Delta and Kappa (B.1.617.2 and B.1.617.1 India). Each of these strains gained advantageous mutations to become a dominant strain, e.g., Iota first discovered November 23, 2020, represented 45% of new cases as of February 7, 2021^1^. Increased transmissibility results from genomic changes such as nonsynonymous mutations in the receptor-binding domain (RBD) of the S-gene (encodes the spike protein) conferring higher binding affinity to host angiotensin-converting enzyme 2 (ACE2) receptors or more efficient cleavage by Transmembrane Serine Protease 2 (TMPRSS2) and subsequently, viral entry^2,3^. Mutations could also lead to vaccine breakthrough^4^. The spike protein’s RBD is immunodominant^5^, targeted by convalescent sera and vaccine-elicited antibodies (Pfizer BNT162b2^6^), though evidence suggest substantial role of the amino-terminal domain (NTD). Mutations in the RBD therefore pose a risk of allowing immune evasion to one or more of the current vaccines^4^. The Kappa and Delta variants emerged from the Indian state of Maharashtra in December 2020, contributing to a resurgence of cases in the country, representing 70% of daily new cases on May 2, 2021^7^. The B.1.617 lineage is now widely circulating in over 50 countries based on viral sequence data and is classified as a variant of concern by the CDC. The Kappa and Delta variant lineages are defined by 7 and 8 nonsynonymous mutations in the S protein respectively (Figure 1).

**Figure 1.**
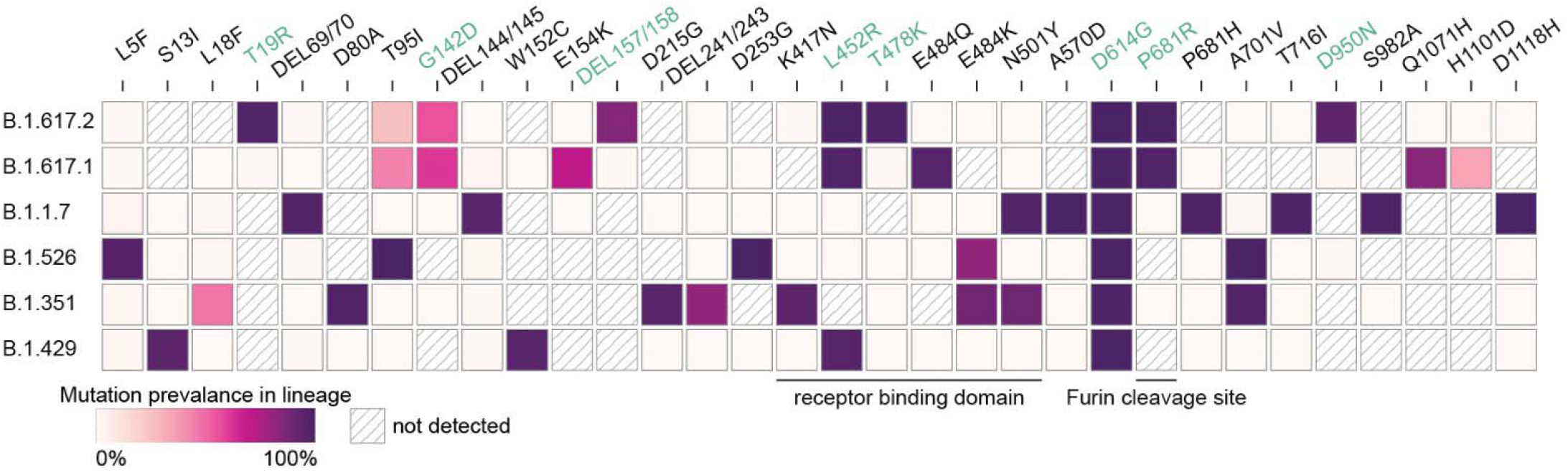
Spike protein mutation prevalence in SARS-CoV-2 variant lineages. Only mutations found in greater than 50% in at least one lineage are displayed. Green text denotes a mutation found in all patients in this report. Figure modified from Outbreak.info^5^

Emergent data suggests partial immunity to the Kappa variant, as convalescent sera and vaccine-elicited (Pfizer BNT162b2 and Moderna mRNA-1273) antibodies show a 2.3- and 4-fold reduction in neutralization in vitro respectively (noting that this study used protein-pseudotyped lentiviruses lacking the T478K mutation found in Delta variant)^8^. A test negative case control study estimated the effectiveness of vaccination (two weeks post second vaccination) against symptomatic disease by Delta variant to be as high as 88% for Pfizer BNT162b2 in the UK (compared to 93% for Alpha variant)^9^.

Here we describe a transmission of a Delta variant containing SARS-CoV-2 strain, between family members associated with events surrounding a wedding with 92 attendees, near Houston, Texas. Attendance required guests be fully vaccinated and took place outdoors in a large, open-air tent. To date, 6 individuals have tested positive for SARS-CoV-2, all patients were symptomatic, one patient severely enough to receive monoclonal antibody infusion treatment (Regeneron Pharmaceuticals Inc.) and one patient has died. Encounter timings and viral sequence similarities suggest the strain containing the Delta variant was transmitted to wedding guests from two patients travelling from India. With no history of vaccine failure in these patients, our observations suggest these are true cases of vaccine breakthrough, mediated by the Delta variant.

## Results

In early April, 2021, Patient 0a, a man with no comorbidities, and Patient 0b, a woman with diabetes, travelled from India to attend a wedding in Texas (designated 0a and 0b due to difficulty establishing true patient 0). Both tested negative for SARS-CoV-2 by qPCR as part of the pre-flight criteria. Formal wedding events were held outdoors and in a large open-air tent and attendance required full vaccination (Patient 0a and 0b travelled to Houston 10 days after their second doses of Covaxin BBV152, Table 1). Patients 1-5 confirmed having close encounters with Patient 0a and 0b at the wedding. Events were attended by fewer than 100 guests.

**Table 1.** Patient demographics, vaccine history, and symptoms

| Sample | Sex | Age | Height (cm) | Weight (kg) | Vaccine received | Symptoms | Comorbidities† | N1 qPCR Ct |
| --- | --- | --- | --- | --- | --- | --- | --- | --- |
| Patient 0a | Male | 67-69 | 177.8 | 81.6 | Covaxin BBV152 | Fever, cough, body aches, fatigue, loss of taste and/or smell, Shortness of breath at rest, Shortness of breath with activity | None | 29 |
| Patient 0b | Female | 65-70 | 152.4 | 53 | Covaxin BBV152 | Fever, cough, body aches, fatigue, loss of taste and/or smell, Shortness of breath at rest, Shortness of breath with activity | Diabetes | 27 |
| Patient 1 | Male | 60-66 | 172 | 68 | Pfizer BNT162b2 | Cough, fatigue | None | 17 |
| Patient 2 | Male | 59-64 | 170.1 | 77 | Pfizer BNT162b2 | Fever, cough, fatigue | Hypertension | 24 |
| Patient 3 | Female | 51-56 | 155 | 63.5 | Moderna mRNA-1273 | Fever, cough, body aches, fatigue, loss of taste and/or smell | Overweight | 25 |
| Patient 4 | Female | 51-56 | 162.6 | 88.5 | Moderna mRNA-1273 | Fatigue, loss of taste and/or smell | Overweight | 22 |

† - As defined by the Centers for Disease Control and Prevention

On the evening of the first night of the wedding, Patient 0b complained of fatigue but associated it with diabetes and jet lag. Patient 0a developed a cough two days after the wedding and both him and Patient 0b developed a fever three days after. Patient 0a and 0b tested positive for SARS-CoV-2 by nasal swab qPCR 4 days after the wedding at a third-party site. Patient 0a’s symptoms progressed over the following days and was admitted to the hospital on day 6 post wedding. He was transferred to Baylor St. Luke’s hospital in the Texas Medical Center with worsening symptoms. A month after the wedding, Patient 0a died from complications of COVID-19.

Four other guests have tested positive for SARS-CoV-2 following confirmed interactions with Patient 0a and 0b. All positive patients received Pfizer BNT162b2, Moderna mRNA-1273, or Covaxin BBV152 (Table 1). Six of these have experienced symptoms of COVID-19 (Table 1). Patient 1, who received the Pfizer BNT162b2 vaccine developed severe symptoms and was admitted to Baylor St. Luke’s hospital for monoclonal antibody infusion treatment (Regeneron Pharmaceuticals Inc.) on ten days after the wedding. The density of vaccine breakthrough resulting in COVID-19 symptoms suggested the patients were carrying a SARS-CoV-2 variant. To characterize the variant, total RNA was extracted from nasopharyngeal swabs of each of the 6 patients. All positive for SARS-CoV-2 Wuhan-Hu-1 and negative for Alpha variant by qPCR (Table 1). Human RNase P (RP) gene control values suggested sampling of patients and RNA isolation were performed optimally. Amplicon libraries were successfully prepared from all 6 qPCR positive samples (N1 Ct value 17 – 29), with 900,754 – 2,381,756 pass-filter reads generated using Swift Biosciences Sarscov2 analysis pipeline (sTable 1). Median sequence coverage ranged from 2085x to 12,932x with >99.7% of the genome covered at 40x or greater. All 6 samples were identified as the SARS-CoV-2 Delta variant based on the presence of the 10 mutations listed by the CDC’s ‘Selected Characteristics of SARS-CoV-2 Variants of Interest’. These mutations, located on the S protein were T19R, (G142D), 156del, 157del, R158G, L452R, T478K, D614G, P681R, D950N. 156del, 157del, R158G are annotated as a single mutation (S:GAGTTCA22028G:Glu156_Arg158delinsGly) due to their proximity (Figure 1). Phylogenetic analysis places each patient sample in a subclade of the Delta variant (white box, Figure 2).

**Figure 2.**
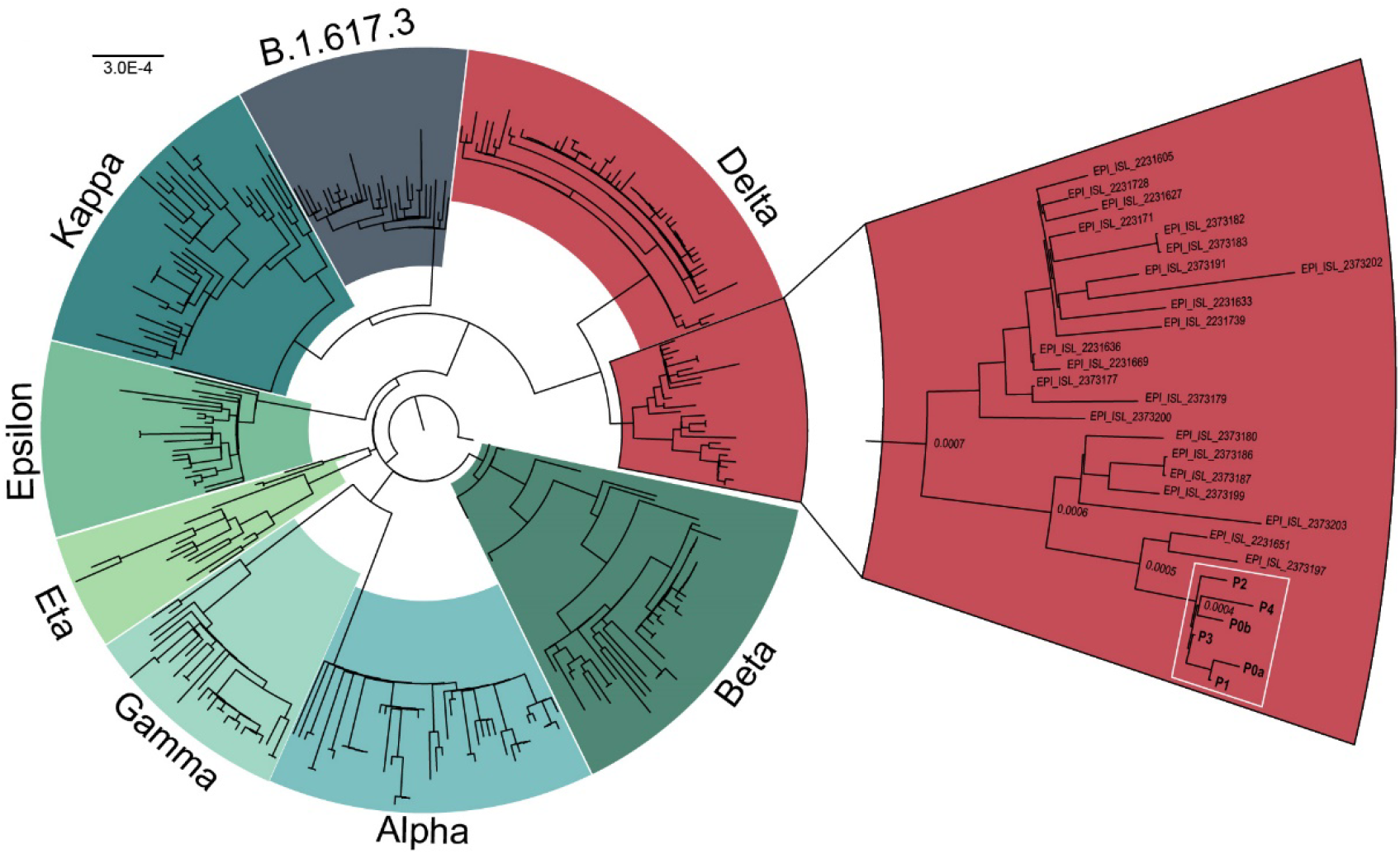
Phylogenetic analysis of SARS-CoV-2 variants. All patients (white box) cluster in a sub clade of the Delta variant (red). Sequences obtained from GISAID.

## Discussion

Ending the current SARS-CoV-2 pandemic requires limiting the spread through continued vigilance of masking, social distancing, and vaccination. Variants emerge from areas experiencing uncontrolled viral spread and display increased transmissibility due to mutations in the spike protein. These mutations may occur in the antigenic region of the RBD, altering binding sites for vaccine-elicited antibodies. Mutations such as the ones found in the Kappa variant, provide partial resistance to antibody neutralization (Pfizer BNT162b2, Moderna mRNA-1273, Regeneron^8^, and Covaxin BBV152^10^), likely due to changes in epitope sequence. Vaccine breakthrough by highly transmissible variants (Delta variant up to 50% more transmissible than Alpha variant, SAGE) could lead to significant setbacks in pandemic control efforts, requiring renewed social distancing and masking efforts. Significant vaccine breakthrough could necessitate vaccine boosters or targeted lockdowns to reduce spread of infection. An analysis of spike protein epitopes found several antigenic regions (IDa-IDi, Zhange et al.). Delta variant spike protein contains mutations in three of these regions (450–469 IDf, 480–499 IDg, and 522–646 IDh, Figure 1) possibly resulting in decreased neutralization by vaccine-elicited antibodies.

According to the cases presented in this study, antibodies elicited in patients receiving Pfizer BNT162b2, Moderna mRNA-1273, and Covaxin BBV152 may provide decreased immunity to the Delta variant. It is possible that some individuals in this study failed to produce an effective immune response to their immunization, however, none of the patients had a history of vaccine failure.

Our observations support continued efforts to generate SARS-CoV-2 genomic sequences from positive patient samples, in order to identify possible vaccine breakthrough mutations. The continued effectiveness of vaccine-elicited antibodies towards SARS-CoV-2 variants highlights the importance of vaccination efforts. Partial efficacy of current vaccines (70% at 75% coverage) is theoretically sufficient to stop a pandemic^11^. Slowing the spread could prevent emergence of future variants, hastening the end of this pandemic.

## Materials and Methods

### Specimen collection and ethical considerations

All individuals were initially tested by third party SARS-CoV-2 testing sites. To confirm, nasopharyngeal swabs were collected by a physician or nurse as close to the first positive test as possible. Samples were submitted to the Alkek Center for Metagenomics and Microbiome Research for RNA extraction and the Human Genome Sequencing Center for qPCR confirmation. Protocols for collection, qPCR testing, and whole genome sequencing were approved by the Baylor College of Medicine Institutional Review Board (H-47423).

### cDNA synthesis and amplicon libraries

RNA extracted from nasopharyngeal swabs of six individuals that tested positive for SARS-COV-2 using qPCR was converted to 1st strand cDNA using SuperScript™ IV First-Strand Synthesis System (Thermo Fisher, Cat. No. 18091050). The 1st strand cDNA reaction was performed starting with 10 µl of total RNA in 25 µl reaction mix, which was incubated at 23°C for 10 minutes followed by 50°C for 50 minutes. The resulting 1^st^ strand cDNA was then diluted with DEPC treated water, where for two samples with Ct<24 cDNA was diluted 20 times and for the remaining four samples, with Ct>24, cDNA was diluted 2 times, respectively. This diluted 1^st^ strand cDNA (10 ul) was used as input for amplification of the SARS-CoV-2 viral genome, using the SARS-CoV-2 Additional Genome Coverage Panel (Cat#COVG1V2-96). This panel was designed against SARS-CoV-2 Wuhan-Hu-1 strain (NC_045512.2) and has 345 amplicons of 116-255 bp (average 150 bp) that cover 99.7% (29,828 of 29,903 total bases) of the genome. These amplicons come in a single tube, and the workflow involves two rounds of PCR, a multiplex PCR (4 + 18 cycles) and the indexing PCR (9 cycles) to generate sequence ready libraries. The reaction mixes and the thermocycler conditions were performed according to the Swift Normalase® Amplicon Panels (SNAP) Workflow. Libraries were barcoded with 8bp unique dual indices at the Indexing PCR. For library normalization, the 2 nM Normalase I protocol was performed on libraries individually, followed by pooling 5 µl of each post-Normalase I library to perform Normalase II reaction, which results in sequence ready library pool. Before sequencing, the normalized library pool concentration was measured using qPCR with KAPA Library Quantification Kits (Roche, KK4835, 07960204001).

### Illumina Sequencing

The pooled SARS-CoV-2 amplicon libraries were sequenced on Illumina NovaSeq 6000 S4 flowcell to generate 2×150bp reads.

### Swift amplicon data analysis

Swift amplicon data were analyzed using Swift Biosciences Sarscov2 analysis pipeline (https://github.com/swiftbiosciences/sarscov2analysis_docker) with minimum read coverage depth of 3. The GATK variants were next filtered with allele fraction >=80, and min read depth 30x^12,13^. Swift analysis pipeline produced variant vcf file, consensus genome, pangolin lineage and Nextclade assignment (https://clades.nextstrain.org/). Variant vcf from Swift amplicon data was also annotated using SnpEff^13^.

### Phylogenetic Analysis

Sequences for the designated variants of concern and variants of interest by centers for disease control (CDC), was downloaded from GISAID on 2^nd^ June 2021^15^. All samples downloaded from GISAID, were analyzed using Pangolin V3.0.3 with pangoLEARN 2021-05-27^16^ to ensure that the variant designation assigned by GISAID is accurate. Global alignment of 334 sequences including the sequences from the current study was done using MAFFT v7.480^17^. Maximum-likelihood phylogenetic tree with boot strap (5,000) was generated using IQ-Tree V2.1.2^18,19^. Annotation and visualization of the tree was carried out by using FigTree v1.4.4 (http://tree.bio.ed.ac.uk/software/figtree/). Clades were labeled with the WHO nomenclature.

## Data Availability

Data will be available on NCBI and GISAID.

## Acknowledgments

Part of this work was supported by the National Institute of Allergy and Infectious Diseases (Grant#1U19AI144297). The authors are grateful to the production teams at HGSC for data generation. We also thank Swift Biosciences for their input for amplicon sequencing. We gratefully acknowledge the authors and the originating and submitting laboratories of the sequences from the GISAID hCov-19 Database on which our research was based. See supplemental acknowledgment for details.

---

**Supplementary Figure 1**. Mutation metrics from sequencing results for all 6 samples. Figure generated using Coronapp^9^

**Supplementary table 1.**
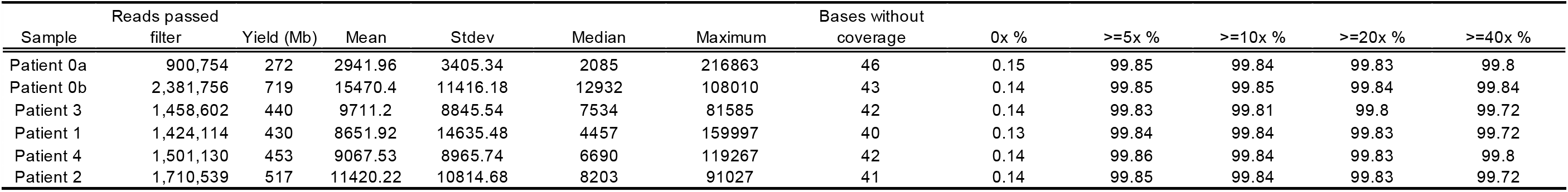
Swift Amplicon Normalase Panel sequencing metrics

