## Supplementary material for "Transmission event of SARS-CoV-2 Delta variant reveals multiple vaccine breakthrough infections": GISAID acknowledgement (from another version of this preprint: version 1, 2021-07-04)

We gratefully acknowledge the following Authors from the Originating laboratories responsible for obtaining the specimens, as well as the Submitting laboratories where the genome data were generated and shared via GISAID, on which this research is based.

All Submitters of data may be contacted directly via www.gisaid.org

Authors are sorted alphabetically.

| Accession ID | Originating Laboratory | Submitting Laboratory | Authors |
|---|---|---|---|
| EPI_ISL_2407454, EPI_ISL_2407457, EPI_ISL_2407459, EPI_ISL_2407460, EPI_ISL_2407462, EPI_ISL_2407464, EPI_ISL_2407465, EPI_ISL_2407470, EPI_ISL_2407472, EPI_ISL_2407475, EPI_ISL_2407478, EPI_ISL_2407479, EPI_ISL_2407483, EPI_ISL_2407485, EPI_ISL_2407490, EPI_ISL_2407491, EPI_ISL_2407492, EPI_ISL_2407493, EPI_ISL_2407496, EPI_ISL_2407497, EPI_ISL_2407501, EPI_ISL_2407507, EPI_ISL_2407509, EPI_ISL_2407510, EPI_ISL_2407519, EPI_ISL_2407520, EPI_ISL_2407530, EPI_ISL_2407534, EPI_ISL_2407537, EPI_ISL_2407538, EPI_ISL_2407543, EPI_ISL_2407549, EPI_ISL_2407550, EPI_ISL_2407551, EPI_ISL_2407553, EPI_ISL_2407555, EPI_ISL_2407559, EPI_ISL_2407563, EPI_ISL_2407564, EPI_ISL_2407565, EPI_ISL_2407567, EPI_ISL_2407569, EPI_ISL_2407570, EPI_ISL_2407572, EPI_ISL_2407575, EPI_ISL_2407576, EPI_ISL_2407578, EPI_ISL_2407580, EPI_ISL_2407583 | | | |
| see above | Lighthouse Lab in Glasgow | Wellcome Sanger Institute for the COVID-19 Genomics UK (COG-UK) Consortium | Harper VanSteenhouse, Yumi Kasai, David Gray, Carol Clugston, Anna Dominiczak and Alex Alderton, Roberto Amato, Jeffrey Barrett, Sonia Goncalves, Ewan Harrison, David K. Jackson, Ian Johnston, Dominic Kwiatkowski, Cordelia Langford, John Sillitoe on behalf of the Wellcome Sanger Institute COVID-19 Surveillance Team |
| EPI_ISL_2408116 | AZ Klina | AZ Klina | Carl Vael - Lynsey Berckmans |

We gratefully acknowledge the following Authors from the Originating laboratories responsible for obtaining the specimens, as well as the Submitting laboratories where the genome data were generated and shared via GISAID, on which this research is based.

All Submitters of data may be contacted directly via www.gisaid.org

Authors are sorted alphabetically.

| Accession ID | Originating Laboratory | Submitting Laboratory | Authors |
|---|---|---|---|
| EPI_ISL_1046783 | SARS-CoV-2 testing team, National Institute of Infectious Diseases | Pathogen Genomics Center, National Institute of Infectious Diseases | Tsuyoshi Sekizuka, Kentaro Itokawa, Rina Tanaka, Masanori Hashino, Eri Nakayama, Shigeru Tajima, Chang-Kweng Lim, Makoto Kuroda |
| EPI_ISL_1073563 | Israel Central Virology laboratory | Israel National Consortium for SARS-CoV-2 sequencing | Neta Zuckerman, Efrat Dahan Bucris, Michal Mandelboim, Dana Bar-Ilan, Oran Erster, Tzvia Mann, Omer Murik, David A. Zeevi, Assaf Rokney, Joseph Jaffe, Eva Nachum, Maya Davidovich Cohen, Ephraim Fass, Gal Zizelski Valenci, Mor Rubinstein, Efrat Rorman, Israel Nissan, Efrat Glick-Saar, Omri Nayshool, Gideon Rechavi, Ella Mendelson, Orna Mor |
| EPI_ISL_1096140 | Cambodian National Public Health Laboratory, National Institute of Public Health | Virology Unit, Institut Pasteur du Cambodge | Sokhoun Yann, Ly Sovann, Kraing Sidonn, Yi Sengdoeurn, Chin Savuth, Chau Darapheak, Veasna Duong, Erik A Karlsson |
| EPI_ISL_1198825 | SARS-CoV-2 testing team, National Institute of Infectious Diseases | Pathogen Genomics Center, National Institute of Infectious Diseases | Tsuyoshi Sekizuka, Kentaro Itokawa, Rina Tanaka, Masanori Hashino, Daisuke Kobayashi, Kento Fukano, Hussein H. Aly, Takanobu Kato, Makoto Kuroda |
| EPI_ISL_1209408, EPI_ISL_1209410, EPI_ISL_1209411, EPI_ISL_1209413, EPI_ISL_1209414, EPI_ISL_1209417, EPI_ISL_1209418, EPI_ISL_1209419, EPI_ISL_1209420, EPI_ISL_1209421, EPI_ISL_1209422, EPI_ISL_1209441, EPI_ISL_1209442 | | | |
| see above | Division of Emerging Infectious Diseases, Bureau of Infectious Diseases Diagnosis Control, Korea Disease Control and Prevention Agency | Division of Emerging Infectious Diseases, Bureau of Infectious Diseases Diagnosis Control, Korea Disease Control and Prevention Agency | Ae Kyung Park, Il-Hwan Kim, Heui Man Kim, Jeong-Min Kim, Chae Young Lee, Jin Sun No, Namjoo Lee, Sang Hee Woo, Eun-Jin Kim |
| EPI_ISL_1358467 | Israel Central Virology laboratory | Israel National Consortium for SARS-CoV-2 sequencing | Neta Zuckerman, Efrat Dahan Bucris, Michal Mandelboim, Dana Bar-Ilan, Oran Erster, Tzvia Mann, Omer Murik, David A. Zeevi, Assaf Rokney, Joseph Jaffe, Eva Nachum, Maya Davidovich Cohen, Ephraim Fass, Gal Zizelski Valenci, Mor Rubinstein, Efrat Rorman, Israel Nissan, Efrat Glick-Saar, Omri Nayshool, Gideon Rechavi, Ella Mendelson, Orna Mor |
| EPI_ISL_1419875, EPI_ISL_1419876 | INSACOG-WB | National Institute of Biomedical Genomics - INSACOG | Arindam Maitra, Bhaswati Bandyopadhyay, Nidhan Kumar Biswas, Tamal Ghosh, Sreedhar Chinnaswamy, Ajay Chakraborti, Saumitra Das |
| EPI_ISL_1426779 | Pathogen Genomics Center, National Institute of Infectious Diseases | Pathogen Genomics Center, National Institute of Infectious Diseases | Tsuyoshi Sekizuka, Kentaro Itokawa, Rina Tanaka, Masanori Hashino, Makoto Kuroda |
| EPI_ISL_1477002 | National Public Health Laboratory, National Centre for Infectious Diseases | National Public Health Laboratory, National Centre for Infectious Diseases | Tze Minn Mak, Zhenyang Zhou, Grace Jie Yin Ngan, Royce Ang, Lin Cui, Raymond Tzer Pin Lin |
| EPI_ISL_1663337 | PRMMCCH, Baripada | Institute of Life Sciences - INSACOG | Sunil K. Raghav, Safal Walia, Arup Ghosh, Atimukta Jha, Amol M. Kanampalliwar, Shifu Aggarwal, Rupesh Dash, Rajeeb Swain, Punit Prasad, INSACOG Consortium, Ajay Parida |
| EPI_ISL_1763797 | Israel Central Virology laboratory | Israel National Consortium for SARS-CoV-2 sequencing | Neta Zuckerman, Efrat Dahan Bucris, Michal Mandelboim, Dana Bar-Ilan, Oran Erster, Tzvia Mann, Omer Murik, David A. Zeevi, Assaf Rokney, Joseph Jaffe, Eva Nachum, Maya Davidovich Cohen, Ephraim Fass, Gal Zizelski Valenci, Mor Rubinstein, Efrat Rorman, Israel Nissan, Efrat Glick-Saar, Omri Nayshool, Gideon Rechavi, Ella Mendelson, Orna Mor |
| EPI_ISL_1790107, EPI_ISL_1790108 | Centre Pasteur du Cameroun | Institut Pasteur de Dakar | Njouom Richard, Diagne Moussa Moïse, Dia Ndongo, Diallo Amadou, Sankhe Safietou, Diop Mamadou, Ndiaye Ndack, Loucoubar Cheikh, Carniel Elisabeth, Faye Ousmane, Sall Amadou Alpha |
| EPI_ISL_1927839 | SARS-CoV-2 testing team, National Institute of Infectious Diseases | Pathogen Genomics Center, National Institute of Infectious Diseases | Tsuyoshi Sekizuka, Kentaro Itokawa, Rina Tanaka, Masanori Hashino, Koichi Ishikawa, Sayuri Seki, Shigeru Kusagawa, Makoto Kuroda |
| EPI_ISL_1927845 | SARS-CoV-2 testing team, National Institute of Infectious Diseases | Pathogen Genomics Center, National Institute of Infectious Diseases | Tsuyoshi Sekizuka, Kentaro Itokawa, Rina Tanaka, Masanori Hashino, Nozomu Hanaoka, Masumichi Saito, Naomi Nojiri, Hazuka Y Furihata, Sana Uchikoba, Tsuguto Fujimoto, Makoto Kuroda |
| EPI_ISL_2083252 | Israel Central Virology laboratory | Israel National Consortium for SARS-CoV-2 sequencing | Neta Zuckerman, Efrat Dahan Bucris, Michal Mandelboim, Dana Bar-Ilan, Oran Erster, Tzvia Mann, Omer Murik, David A. Zeevi, Assaf Rokney, Joseph Jaffe, Eva Nachum, Maya Davidovich Cohen, Ephraim Fass, Gal Zizelski Valenci, Mor Rubinstein, Efrat Rorman, Israel Nissan, Efrat Glick-Saar, Omri Nayshool, Gideon Rechavi, Ella Mendelson, Orna Mor |
| EPI_ISL_2158615 | Department of Veterinary Science, National Institute of Infectious Diseases | Department of Veterinary Science, National Institute of Infectious Diseases | Yudai Kuroda, Tsukasa Yamamoto, Keita Ishijima, Souichi Yamada, Shuetsu Fukushi, Ken Maeda |
| EPI_ISL_768628 | National Public Health Laboratory, National Centre for Infectious Diseases | National Public Health Laboratory, National Centre for Infectious Diseases | Tze Minn Mak, Sophie Octavia, Zhenyang Zhou, Lin Cui, Raymond Tzer Pin Lin |
| EPI_ISL_768715, EPI_ISL_779199, EPI_ISL_779200, EPI_ISL_779215, EPI_ISL_779234, EPI_ISL_779665, EPI_ISL_779690 | Pathogen Genomics Center, National Institute of Infectious Diseases | Pathogen Genomics Center, National Institute of Infectious Diseases | Tsuyoshi Sekizuka, Kentaro Itokawa, Rina Tanaka, Masanori Hashino, Makoto Kuroda |
| EPI_ISL_804188 | Israel Central Virology laboratory | Israel National Consortium for SARS-CoV-2 sequencing | Neta Zuckerman, Efrat Dahan Bucris, Michal Mandelboim, Dana Bar-Ilan, Oran Erster, Tzvia Mann, Omer Murik, David A. Zeevi, Assaf Rokney, Joseph Jaffe, Eva Nachum, Maya Davidovich Cohen, Ephraim Fass, Gal Zizelski Valenci, Mor Rubinstein, Efrat Rorman, Israel Nissan, Efrat Glick-Saar, Omri Nayshool, Gideon Rechavi, Ella Mendelson, Orna Mor |
| EPI_ISL_825411, EPI_ISL_825421, EPI_ISL_825427, EPI_ISL_860106, EPI_ISL_860108, EPI_ISL_860112 | Pathogen Genomics Center, National Institute of Infectious Diseases | Pathogen Genomics Center, National Institute of Infectious Diseases | Tsuyoshi Sekizuka, Kentaro Itokawa, Rina Tanaka, Masanori Hashino, Makoto Kuroda |
| EPI_ISL_944484 | Israel Central Virology laboratory | Israel National Consortium for SARS-CoV-2 sequencing | Neta Zuckerman, Efrat Dahan Bucris, Michal Mandelboim, Dana Bar-Ilan, Oran Erster, Tzvia Mann, Omer Murik, David A. Zeevi, Assaf Rokney, Joseph Jaffe, Eva Nachum, Maya Davidovich Cohen, Ephraim Fass, Gal Zizelski Valenci, Mor Rubinstein, Efrat Rorman, Israel Nissan, Efrat Glick-Saar, Omri Nayshool, Gideon Rechavi, Ella Mendelson, Orna Mor |
| EPI_ISL_995299, EPI_ISL_995302 | National Public Health Laboratory, National Centre for Infectious Diseases | National Public Health Laboratory, National Centre for Infectious Diseases | Tze Minn Mak, Zhenyang Zhou, Lin Cui, Raymond Tzer Pin Lin |

We gratefully acknowledge the following Authors from the Originating laboratories responsible for obtaining the specimens, as well as the Submitting laboratories where the genome data were generated and shared via GISAID, on which this research is based.

All Submitters of data may be contacted directly via www.gisaid.org

Authors are sorted alphabetically.

| Accession ID | Originating Laboratory | Submitting Laboratory | Authors |
|---|---|---|---|
| EPI_ISL_1173247 | Piyavate Hospital | National Institute of Health, Department of Medical Sciences, Ministry of Public Health, Thailand | Pilailuk Okada; Siripaporn Phuygun; Sittiporn Parnmen; Nuttida Thongpramul; Ratana Tacharoenmuang; Pakorn Piromtong; Natchaya Khiadsang; Thanutsapa Thanadachakul; Warawan Wongboot; Sunthareeya Waicharoen. |
| EPI_ISL_1180280 | National Virus Reference Laboratory | National Virus Reference Laboratory | Guerrino Macori, Gabriel Gonzalez, Michael Carr, Zoe Yandle, Charlene Bennett, Jonathan Dean, Seamus Fanning, Cillian F De Gascun |
| EPI_ISL_1469122, EPI_ISL_1469145, EPI_ISL_1469155 | AZDelta | AZDelta | Geert Martens; Dieter De Smet |
| EPI_ISL_1663334 | PRMMCCH, Baripada | Institute of Life Sciences - INSACOG | Sunil K. Raghav, Safal Walia, Arup Ghosh, Atimukta Jha, Amol M. Kanampalliwar, Shifu Aggarwal, Rupesh Dash, Rajeeb Swain, Punit Prasad, INSACOG Consortium, Ajay Parida |
| EPI_ISL_1663494 | Capital hospital, Bhubaneswar | Institute of Life Sciences - INSACOG | Sunil K. Raghav, Safal Walia, Arup Ghosh, Atimukta Jha, Amol M. Kanampalliwar, Shifu Aggarwal, Rupesh Dash, Rajeeb Swain, Punit Prasad, INSACOG Consortium, Ajay Parida |
| EPI_ISL_1695986 | GH COCHIN ST-VINCENT DE PAUL | Department of Virology, Henri Mondor University Hospital, Assistance Publique Hôpitaux de Paris, Université Paris-Est Créteil, INSERM U955 | Christophe Rodriguez, Slim Fourati, Vanessa Demontant, Guillaume Gricourt, Melissa N'Debi, Alexandre Soulier, Elisabeth Trawinski, Jean-Michel Pawlotsky |
| EPI_ISL_1716799 | CSIR-IGIB | CSIR-IGIB | Rajesh Pandey, Janani SV, Ranjeet Maurya, Akshay Kanakan, Neha Jha, Priyanka Mehta, Partha Chattopadhyay, Priti Devi, Shweta Sahni, Azka Khan, Anish Maitra |
| EPI_ISL_1754921, EPI_ISL_1754931 | Hôpital Cochin | Department of Virology, Henri Mondor University Hospital, Assistance Publique Hôpitaux de Paris, Université Paris-Est Créteil, INSERM U955 | Christophe Rodriguez, Slim Fourati, Vanessa Demontant, Guillaume Gricourt, Melissa N'Debi, Alexandre Soulier, Elisabeth Trawinski, Jean-Michel Pawlotsky |
| EPI_ISL_1754998 | Hôpital Avicenne | Department of Virology, Henri Mondor University Hospital, Assistance Publique Hôpitaux de Paris, Université Paris-Est Créteil, INSERM U955 | Christophe Rodriguez, Slim Fourati, Vanessa Demontant, Guillaume Gricourt, Melissa N'Debi, Alexandre Soulier, Elisabeth Trawinski, Jean-Michel Pawlotsky |
| EPI_ISL_1755014 | CH.INTERCOMMUNAL DE CRETEIL | Department of Virology, Henri Mondor University Hospital, Assistance Publique Hôpitaux de Paris, Université Paris-Est Créteil, INSERM U955 | Christophe Rodriguez, Slim Fourati, Vanessa Demontant, Guillaume Gricourt, Melissa N'Debi, Alexandre Soulier, Elisabeth Trawinski, Jean-Michel Pawlotsky |
| EPI_ISL_1755030 | GH A.CHENEVIER-H.MONDOR | Department of Virology, Henri Mondor University Hospital, Assistance Publique Hôpitaux de Paris, Université Paris-Est Créteil, INSERM U955 | Christophe Rodriguez, Slim Fourati, Vanessa Demontant, Guillaume Gricourt, Melissa N'Debi, Alexandre Soulier, Elisabeth Trawinski, Jean-Michel Pawlotsky |
| EPI_ISL_1756563, EPI_ISL_1756612, EPI_ISL_1756631, EPI_ISL_1756637 | Biogroup Bio Lam-LCD Saint-Denis | Department of Virology, Henri Mondor University Hospital, Assistance Publique Hôpitaux de Paris, Université Paris-Est Créteil, INSERM U955 | Christophe Rodriguez, Slim Fourati, Vanessa Demontant, Guillaume Gricourt, Melissa N'Debi, Alexandre Soulier, Elisabeth Trawinski, Jean-Michel Pawlotsky |
| EPI_ISL_1760146 | Ministry of Health Turkey | Ministry of Health Turkey | Fatma Bayrakdar, Yasemin Cosgun, Suleyman Yalcin, Gulay Korukluoglu |
| EPI_ISL_1817591, EPI_ISL_1817648 | Servicio Microbiología Hospital La Paz | Servicio Microbiología Hospital La Paz | Fernando Lázaro, Rubén Cáceres, Jesús Mingorance Cruz, Elie Dahdouh |
| EPI_ISL_1838719, EPI_ISL_1838748 | CSIR-Centre for Cellular and Molecular Biology | CSIR-Centre for Cellular and Molecular Biology-INSACOG | Payel Mukherjee,Lamuk Zaveri,Tulasi Nagabandi, Ara Sreenivas,Shreekant Verma, Amareshwar Vodapalli ,Blessy B John,Viswagithe S L,B Himasri,Valli Nagalakshmi Undamatla,Onkar Kulkarni,Sofia Banu,Archana Bharadwaj Siva,Sharath Chandra Thota,Karthik Bharadwaj Tallapaka,Rakesh K Mishra,Divya Tej Sowpati |
| EPI_ISL_2033624 | Swedish national genomic surveillance program of SARS-CoV-2 | The Public Health Agency of Sweden | Maximilian Riess, Maria Lind Karlberg, Alma Brolund, Swedish national genomic surveillance program of SARS-CoV-2 |
| EPI_ISL_2156972 | Mapmygenome | CSIR-Centre for Cellular and Molecular Biology-INSACOG | Ara Sreenivas,Shreekant Verma,Tulasi Nagabandi, Lamuk Zaveri, Amareshwar Vodapalli ,Blessy B John,Viswagithe S L,B Himasri,Valli Nagalakshmi Undamatla,Payel Mukherjee,Sofia Banu,Archana Bharadwaj Siva,Sharath Chandra Thota,Karthik Bharadwaj Tallapaka,Rakesh K Mishra,Divya Tej Sowpati |
| EPI_ISL_2179748, EPI_ISL_2179749 | Servicio Microbiología Hospital La Paz | Servicio Microbiología Hospital La Paz | Fernando Lázaro, Rubén Cáceres, Jesús Mingorance Cruz, Elie Dahdouh |
| EPI_ISL_2373988 | Laboratoire Virologie Saint Louis APHP | Laboratoire Virologie Saint Louis APHP | Maud Salmona, Marie Laure Chaix, Severine Mercier Delarue, Marie Laure Néré, Linda Feghoul, Jérome Le Goff, Constance Delauguerre, Sophia Achaibou |
| EPI_ISL_2379487 | Dr S Raju, Director of Public Heath and Preventive Medicine | inStem NCBS | Uma Ramakrishnan Dasaradhi Palakodeti Aswin SaiNarain |

We gratefully acknowledge the following Authors from the Originating laboratories responsible for obtaining the specimens, as well as the Submitting laboratories where the genome data were generated and shared via GISAID, on which this research is based.

All Submitters of data may be contacted directly via www.gisaid.org

Authors are sorted alphabetically.

| Accession ID | Originating Laboratory | Submitting Laboratory | Authors |
|---|---|---|---|
| EPI_ISL_2407502, EPI_ISL_2407503, EPI_ISL_2407505, EPI_ISL_2407506, EPI_ISL_2407508, EPI_ISL_2407511, EPI_ISL_2407512, EPI_ISL_2407513, EPI_ISL_2407515, EPI_ISL_2407516, EPI_ISL_2407517, EPI_ISL_2407521, EPI_ISL_2407522, EPI_ISL_2407523, EPI_ISL_2407524, EPI_ISL_2407525, EPI_ISL_2407526, EPI_ISL_2407527, EPI_ISL_2407528, EPI_ISL_2407529, EPI_ISL_2407531, EPI_ISL_2407532, EPI_ISL_2407535, EPI_ISL_2407536, EPI_ISL_2407539, EPI_ISL_2407542, EPI_ISL_2407544, EPI_ISL_2407545, EPI_ISL_2407546, EPI_ISL_2407547, EPI_ISL_2407548, EPI_ISL_2407552, EPI_ISL_2407554, EPI_ISL_2407556, EPI_ISL_2407560, EPI_ISL_2407561, EPI_ISL_2407562, EPI_ISL_2407566, EPI_ISL_2407568, EPI_ISL_2407571, EPI_ISL_2407573, EPI_ISL_2407574, EPI_ISL_2407577, EPI_ISL_2407579, EPI_ISL_2407581, EPI_ISL_2407582, EPI_ISL_2407584, EPI_ISL_2407585, EPI_ISL_2407587, EPI_ISL_2407589 | | | |
| see above | Lighthouse Lab in Glasgow | Wellcome Sanger Institute for the COVID-19 Genomics UK (COG-UK) Consortium | Harper VanSteenhouse, Yumi Kasai, David Gray, Carol Clugston, Anna Dominiczak and Alex Alderton, Roberto Amato, Jeffrey Barrett, Sonia Goncalves, Ewan Harrison, David K. Jackson, Ian Johnston, Dominic Kwiatkowski, Cordelia Langford, John Sillitoe on behalf of the Wellcome Sanger Institute COVID-19 Surveillance Team |

We gratefully acknowledge the following Authors from the Originating laboratories responsible for obtaining the specimens, as well as the Submitting laboratories where the genome data were generated and shared via GISAID, on which this research is based.

All Submitters of data may be contacted directly via www.gisaid.org

Authors are sorted alphabetically.

| Accession ID | Originating Laboratory | Submitting Laboratory | Authors |
|---|---|---|---|
| EPI_ISL_2231598, EPI_ISL_2231605, EPI_ISL_2231627 | AP SSO | CSIR-Centre for Cellular and Molecular Biology-INSACOG | B Himasri,Valli Nagalakshmi Undamatla,Lamuk Zaveri,Tulasi Nagabandi, Ara Sreenivas,Shreekant Verma, Amareshwar Vodapalli ,Blessy B John,Viswagithe S L,Payel Mukherjee,Sofia Banu,Archana Bharadwaj Siva,Sharath Chandra Thota,Karthik Bharadwaj Tallapaka,Rakesh K Mishra,Divya Tej Sowpati |
| EPI_ISL_2231633, EPI_ISL_2231636, EPI_ISL_2231640, EPI_ISL_2231650, EPI_ISL_2231651, EPI_ISL_2231654, EPI_ISL_2231669, EPI_ISL_2231695 | AP SSO | CSIR-Centre for Cellular and Molecular Biology-INSACOG | Tulasi Nagabandi, Lamuk Zaveri,Ara Sreenivas,Shreekant Verma, Amareshwar Vodapalli ,Blessy B John,Viswagithe S L,B Himasri,Valli Nagalakshmi Undamatla,Payel Mukherjee,Sofia Banu,Archana Bharadwaj Siva,Sharath Chandra Thota,Karthik Bharadwaj Tallapaka,Rakesh K Mishra,Divya Tej Sowpati |
| EPI_ISL_2231711 | AP SSO | CSIR-Centre for Cellular and Molecular Biology-INSACOG | B Himasri,Valli Nagalakshmi Undamatla,Lamuk Zaveri,Tulasi Nagabandi, Ara Sreenivas,Shreekant Verma, Amareshwar Vodapalli ,Blessy B John,Viswagithe S L,Payel Mukherjee,Sofia Banu,Archana Bharadwaj Siva,Sharath Chandra Thota,Karthik Bharadwaj Tallapaka,Rakesh K Mishra,Divya Tej Sowpati |
| EPI_ISL_2231712, EPI_ISL_2231728 | AP SSO | CSIR-Centre for Cellular and Molecular Biology-INSACOG | Tulasi Nagabandi, Lamuk Zaveri,Ara Sreenivas,Shreekant Verma, Amareshwar Vodapalli ,Blessy B John,Viswagithe S L,B Himasri,Valli Nagalakshmi Undamatla,Payel Mukherjee,Sofia Banu,Archana Bharadwaj Siva,Sharath Chandra Thota,Karthik Bharadwaj Tallapaka,Rakesh K Mishra,Divya Tej Sowpati |
| EPI_ISL_2231739 | AP SSO | CSIR-Centre for Cellular and Molecular Biology-INSACOG | B Himasri,Valli Nagalakshmi Undamatla,Lamuk Zaveri,Tulasi Nagabandi, Ara Sreenivas,Shreekant Verma, Amareshwar Vodapalli ,Blessy B John,Viswagithe S L,Payel Mukherjee,Sofia Banu,Archana Bharadwaj Siva,Sharath Chandra Thota,Karthik Bharadwaj Tallapaka,Rakesh K Mishra,Divya Tej Sowpati |
| EPI_ISL_2373177, EPI_ISL_2373178, EPI_ISL_2373179, EPI_ISL_2373180, EPI_ISL_2373182 | AP SSO | CSIR-Centre for Cellular and Molecular Biology-INSACOG | Tulasi Nagabandi, Lamuk Zaveri,Ara Sreenivas,Shreekant Verma, Amareshwar Vodapalli ,Blessy B John,Viswagithe S L,B Himasri,Valli Nagalakshmi Undamatla,Payel Mukherjee,Sofia Banu,Archana Bharadwaj Siva,Sharath Chandra Thota,Karthik Bharadwaj Tallapaka,Rakesh K Mishra,Divya Tej Sowpati |
| EPI_ISL_2373183, EPI_ISL_2373185, EPI_ISL_2373186, EPI_ISL_2373187, EPI_ISL_2373188, EPI_ISL_2373189, EPI_ISL_2373190, EPI_ISL_2373191, EPI_ISL_2373193, EPI_ISL_2373197, EPI_ISL_2373198, EPI_ISL_2373199, EPI_ISL_2373200, EPI_ISL_2373201, EPI_ISL_2373202, EPI_ISL_2373203, EPI_ISL_2373204 | | | |
| see above | AP SSO | CSIR-Centre for Cellular and Molecular Biology-INSACOG | Blessy B John,Viswagithe S L,Tulasi Nagabandi, Lamuk Zaveri,Ara Sreenivas,Shreekant Verma, Amareshwar Vodapalli ,B Himasri,Valli Nagalakshmi Undamatla,Payel Mukherjee,Sofia Banu,Archana Bharadwaj Siva,Sharath Chandra Thota,Karthik Bharadwaj Tallapaka,Rakesh K Mishra,Divya Tej Sowpati |
| EPI_ISL_2407587 | Lighthouse Lab in Glasgow | Wellcome Sanger Institute for the COVID-19 Genomics UK (COG-UK) Consortium | Harper VanSteenhouse, Yumi Kasai, David Gray, Carol Clugston, Anna Dominiczak and Alex Alderton, Roberto Amato, Jeffrey Barrett, Sonia Goncalves, Ewan Harrison, David K. Jackson, Ian Johnston, Dominic Kwiatkowski, Cordelia Langford, John Sillitoe on behalf of the Wellcome Sanger Institute COVID-19 Surveillance Team |

We gratefully acknowledge the following Authors from the Originating laboratories responsible for obtaining the specimens, as well as the Submitting laboratories where the genome data were generated and shared via GISAID, on which this research is based.

All Submitters of data may be contacted directly via www.gisaid.org

Authors are sorted alphabetically.

| Accession ID | Originating Laboratory | Submitting Laboratory | Authors |
|---|---|---|---|
| EPI_ISL_2405049, EPI_ISL_2405089 | UW Virology Lab | UW Virology Lab | Pavitra Roychoudhury, Hong Xie, Lasata Shrestha, Shah Mohamed Bakhash, Noah R. Baker, Sean Ellis, Saraswathi Sathees, Meei-Li Huang, Keith R Jerome, Alexander Greninger |
| EPI_ISL_2405167 | UCLA Clinical Micro Lab | Los Angeles County PHL | P. Hemarajata et al. |
| EPI_ISL_2405204 | Pomona Valley Hospital Medical Center | Los Angeles County PHL | P. Hemarajata et al. |
| EPI_ISL_2405336 | University Hospitals of Geneva, Laboratory of Virology | HUG, Laboratory of Virology and the Health2030 Genome Center | Samuel Cordey, Ana Rita Goncalves, Laurent Kaiser, Lorenzo Cerutti, Henri Pegeot, Melyssa Elies, Deborah Penet, Keith Harshman, Ioannis Xenarios, Emmanouil Dermitzakis |
| EPI_ISL_2405430, EPI_ISL_2405451, EPI_ISL_2405523, EPI_ISL_2405534, EPI_ISL_2405546, EPI_ISL_2405560, EPI_ISL_2405571, EPI_ISL_2405572, EPI_ISL_2405606, EPI_ISL_2405658, EPI_ISL_2405753, EPI_ISL_2405788, EPI_ISL_2405834, EPI_ISL_2405886, EPI_ISL_2405922, EPI_ISL_2406008, EPI_ISL_2406028, EPI_ISL_2406085, EPI_ISL_2406108, EPI_ISL_2406122, EPI_ISL_2406150, EPI_ISL_2406198, EPI_ISL_2406199, EPI_ISL_2406225, EPI_ISL_2406236, EPI_ISL_2406247, EPI_ISL_2406248, EPI_ISL_2406259, EPI_ISL_2406269, EPI_ISL_2406320, EPI_ISL_2406331, EPI_ISL_2406344, EPI_ISL_2406366, EPI_ISL_2406374, EPI_ISL_2406397, EPI_ISL_2406413, EPI_ISL_2406433, EPI_ISL_2406434, EPI_ISL_2406440, EPI_ISL_2406445, EPI_ISL_2406457 | | | |
| see above | Dutch COVID-19 response team | National Institute for Public Health and the Environment (RIVM) | Adam Meijer, Harry Vennema, Dirk Eggink, Jeroen Cremer, Sharon van den Brink, Bas van der Veer, AnneMarie van den Brandt, Lisa Wijsman, Kim Freriks, Ryanne Jaarsma, Euníce Then, Lynn Aarts, Sanne Bos, Melissa van Tuil, Linda van de Nes, Sjoerd Kuiling, James Groot, Florian Zwagemaker, Dennis Schmitz, Annelies Kroneman, Karim Hajji, Chantal Reusken, on behalf of the national COVID-19 response team |
| EPI_ISL_2406491 | Middlemore Hospital | Institute of Environmental Science and Research (ESR) | Rachel Boyle, SallyAnn Harbison, Olivia Stroeven, Xiaoyun Ren, Matt Storey, Nikki Freed, Muhammad Faisal, Jing Wang, Hermes Perez, Anja Werno, Antje van der Linden, Arlo Upton, Chris Mansell, David Hammer, Dragana Drinkovic, Gary McAuliffe, Hana Sofia Andersson, James Ussher, Jill Sherwood, Josh Freeman, Julia Howard, Juliet Elvy, Mary DeAlmeida, Matt Blakiston, Matthew Rogers, Max Bloomfield, Michael Addidle, Michelle Balm, Sally Roberts, Sarah Jefferies, Sharmini Muttaiyah, Susan Morpeth, Susan Taylor, Timothy Blackmore, Vani Sathyendran, Veronica Playle, Virginia Hope, Erasmus Smit, Lauren Jelly, Olin Silander, Joep de Ligt |
| EPI_ISL_2407652 | Hospital de San Agustín | Laboratorio de Virología HUCA | Sandoval M, Castelló C, Gómez de Oña J, Boga JA, Rojo S, Alvarez-Arguelles ME, Abreu F, Costales I, Perez-Martínez Z, Martín-Rodríguez G, Coto E, Melón S |
| EPI_ISL_2408062, EPI_ISL_2408063 | Usansolo-Galdakao University Hospital | Cruces University Hospital | Ana Gual-de-Torrella, Izaskun Alejo-Cancho, Mikel Gallego, Ana Belén de la Hoz |

We gratefully acknowledge the following Authors from the Originating laboratories responsible for obtaining the specimens, as well as the Submitting laboratories where the genome data were generated and shared via GISAID, on which this research is based.

All Submitters of data may be contacted directly via www.gisaid.org

Authors are sorted alphabetically.

| Accession ID | Originating Laboratory | Submitting Laboratory | Authors |
|---|---|---|---|
| EPI_ISL_2403233 | CHU Pontchaillou | CHU Pontchaillou | GROLHIER Claire, DENOUAL Florent, ETCHEVERRY Amandine, SASSI Mohamed, JAGLINE Steven, FEBREAU Christine, PRONIER Charlotte, GALIBERT Marie Dominique, DE TAYRAC Marie, THIBAULT Vincent |
| EPI_ISL_2403300, EPI_ISL_2403312, EPI_ISL_2403330, EPI_ISL_2403347, EPI_ISL_2403353, EPI_ISL_2403354, EPI_ISL_2403536, EPI_ISL_2403560 | Ministry of Health Turkey | Ministry of Health Turkey | Fatma Bayrakdar, Yasemin Cosgun, Suleyman Yalcin, Gulay Korukluoglu |
| EPI_ISL_2403871, EPI_ISL_2403872, EPI_ISL_2403873, EPI_ISL_2403874, EPI_ISL_2403875, EPI_ISL_2403876, EPI_ISL_2403877, EPI_ISL_2403878, EPI_ISL_2403879, EPI_ISL_2403881, EPI_ISL_2403937, EPI_ISL_2403938, EPI_ISL_2403951, EPI_ISL_2403957, EPI_ISL_2403992, EPI_ISL_2403993, EPI_ISL_2403998, EPI_ISL_2404000, EPI_ISL_2404050, EPI_ISL_2404064, EPI_ISL_2404069, EPI_ISL_2404070, EPI_ISL_2404071, EPI_ISL_2404072, EPI_ISL_2404080, EPI_ISL_2404081, EPI_ISL_2404085, EPI_ISL_2404199, EPI_ISL_2404200, EPI_ISL_2404623 | | | |
| see above | KU Leuven, Rega Institute, Clinical and Epidemiological Virology | KU Leuven, Rega Institute, Clinical and Epidemiological Virology | Tony Wawina-Bokalanga, Bert Vanmechelen, Joan Marti-Carerras, Piet Maes |
| EPI_ISL_2404745 | Oregon State Public Health Laboratory | Oregon State Public Health Laboratory | Rafia Razzaque, Eugene Yeboah, Vanda Makris, Laura Tsaknaridis, John Fontana and Shane Sevey |
| EPI_ISL_2405005, EPI_ISL_2405006, EPI_ISL_2405025 | UW Virology Lab | UW Virology Lab | Pavitra Roychoudhury, Hong Xie, Lasata Shrestha, Shah Mohamed Bakhash, Noah R. Baker, Sean Ellis, Saraswathi Sathees, Meei-Li Huang, Keith R Jerome, Alexander Greninger |
| EPI_ISL_2405473, EPI_ISL_2405485, EPI_ISL_2406058, EPI_ISL_2406090 | Dutch COVID-19 response team | National Institute for Public Health and the Environment (RIVM) | Adam Meijer, Harry Vennema, Dirk Eggink, Jeroen Cremer, Sharon van den Brink, Bas van der Veer, AnneMarie van den Brandt, Lisa Wijsman, Kim Freriks, Ryanne Jaarsma, Euníce Then, Lynn Aarts, Sanne Bos, Melissa van Tuil, Linda van de Nes, Sjoerd Kuiling, James Groot, Florian Zwagemaker, Dennis Schmitz, Annelies Kroneman, Karim Hajji, Chantal Reusken, on behalf of the national COVID-19 response team |
| EPI_ISL_2406492 | Middlemore Hospital | Institute of Environmental Science and Research (ESR) | Rachel Boyle, SallyAnn Harbison, Olivia Stroeven, Xiaoyun Ren, Matt Storey, Nikki Freed, Muhammad Faisal, Jing Wang, Hermes Perez, Anja Werno, Antje van der Linden, Arlo Upton, Chris Mansell, David Hammer, Dragana Drinkovic, Gary McAuliffe, Hana Sofia Andersson, James Ussher, Jill Sherwood, Josh Freeman, Julia Howard, Juliet Elvy, Mary DeAlmeida, Matt Blakiston, Matthew Rogers, Max Bloomfield, Michael Addidle, Michelle Balm, Sally Roberts, Sarah Jefferies, Sharmini Muttaiyah, Susan Morpeth, Susan Taylor, Timothy Blackmore, Vani Sathyendran, Veronica Playle, Virginia Hope, Erasmus Smit, Lauren Jelly, Olin Silander, Joep de Ligt |
| EPI_ISL_2406499, EPI_ISL_2406502 | Laboratoire Virologie Saint Louis APHP | Laboratoire Virologie Saint Louis APHP | Maud Salmona, Marie Laure Chaix, Severine Mercier Delarue, Marie Laure Néré, Linda Feghoul, Jérome Le Goff, Constance Delauguerre, Sophia Achaibou |

We gratefully acknowledge the following Authors from the Originating laboratories responsible for obtaining the specimens, as well as the Submitting laboratories where the genome data were generated and shared via GISAID, on which this research is based.

All Submitters of data may be contacted directly via www.gisaid.org

Authors are sorted alphabetically.

| Accession ID | Originating Laboratory | Submitting Laboratory | Authors |
|---|---|---|---|
| EPI_ISL_1360304 | CSIR-National Environmental Engineering Research Institute | CSIR-Centre for Cellular and Molecular Biology - INSACOG | Onkar Kulkarni, Lamuk Zaveri, Ara Sreenivas, Sofia Banu, Shreekant Verma, Amareshwar Vodapalli , Viswagithe S L, B Himasri, Sharath Chandra Thota, Karthik Bharadwaj Tallapaka, Krishna Khairnar, Rakesh K Mishra,Divya Tej Sowpati |
| EPI_ISL_1360306 | CSIR-National Environmental Engineering Research Institute | CSIR-Centre for Cellular and Molecular Biology - INSACOG | Lamuk Zaveri, Ara Sreenivas, Onkar Kulkarni, Sofia Banu, Shreekant Verma,Amareshwar Vodapalli, Viswagithe S L, B Himasri, Sharath Chandra Thota, Karthik Bharadwaj Tallapaka, Krishna Khairnar, Rakesh K Mishra,Divya Tej Sowpati |
| EPI_ISL_1360316 | CSIR-National Environmental Engineering Research Institute | CSIR-Centre for Cellular and Molecular Biology - INSACOG | Ara Sreenivas, Onkar Kulkarni, Lamuk Zaveri, Sofia Banu, Shreekant Verma, Amareshwar Vodapalli , Viswagithe S L, B Himasri, Sharath Chandra Thota, Karthik Bharadwaj Tallapaka, Krishna Khairnar, Rakesh K Mishra,Divya Tej Sowpati |
| EPI_ISL_1360317, EPI_ISL_1360318 | CSIR-National Environmental Engineering Research Institute | CSIR-Centre for Cellular and Molecular Biology - INSACOG | Sofia Banu, Lamuk Zaveri, Ara Sreenivas, Shreekant Verma, Onkar Kulkarni, Amareshwar Vodapalli ,Viswagithe S L, B Himasri, Sharath Chandra Thota, Karthik Bharadwaj Tallapaka, Krishna Khairnar, Rakesh K Mishra,Divya Tej Sowpati |
| EPI_ISL_1360328 | CSIR-National Environmental Engineering Research Institute | CSIR-Centre for Cellular and Molecular Biology - INSACOG | Onkar Kulkarni, Lamuk Zaveri, Ara Sreenivas, Sofia Banu, Shreekant Verma, Amareshwar Vodapalli , Viswagithe S L, B Himasri, Sharath Chandra Thota, Karthik Bharadwaj Tallapaka, Krishna Khairnar, Rakesh K Mishra,Divya Tej Sowpati |
| EPI_ISL_1360329 | CSIR-National Environmental Engineering Research Institute | CSIR-Centre for Cellular and Molecular Biology - INSACOG | Lamuk Zaveri, Ara Sreenivas, Sofia Banu, Onkar Kulkarni, Shreekant Verma, Amareshwar Vodapalli , Viswagithe S L, B Himasri, Sharath Chandra Thota, Karthik Bharadwaj Tallapaka, Krishna Khairnar, Rakesh K Mishra,Divya Tej Sowpati |
| EPI_ISL_1360330, EPI_ISL_1360338 | CSIR-National Environmental Engineering Research Institute | CSIR-Centre for Cellular and Molecular Biology - INSACOG | Lamuk Zaveri, Ara Sreenivas, Onkar Kulkarni, Sofia Banu, Shreekant Verma,Amareshwar Vodapalli, Viswagithe S L, B Himasri, Sharath Chandra Thota, Karthik Bharadwaj Tallapaka, Krishna Khairnar, Rakesh K Mishra,Divya Tej Sowpati |
| EPI_ISL_1360342 | CSIR-National Environmental Engineering Research Institute | CSIR-Centre for Cellular and Molecular Biology - INSACOG | Sofia Banu, Lamuk Zaveri, Ara Sreenivas, Shreekant Verma, Onkar Kulkarni, Amareshwar Vodapalli ,Viswagithe S L, B Himasri, Sharath Chandra Thota, Karthik Bharadwaj Tallapaka, Krishna Khairnar, Rakesh K Mishra,Divya Tej Sowpati |
| EPI_ISL_1360352 | CSIR-National Environmental Engineering Research Institute | CSIR-Centre for Cellular and Molecular Biology - INSACOG | Onkar Kulkarni, Lamuk Zaveri, Ara Sreenivas, Sofia Banu, Shreekant Verma, Amareshwar Vodapalli , Viswagithe S L, B Himasri, Sharath Chandra Thota, Karthik Bharadwaj Tallapaka, Krishna Khairnar, Rakesh K Mishra,Divya Tej Sowpati |
| EPI_ISL_1360359 | CSIR-National Environmental Engineering Research Institute | CSIR-Centre for Cellular and Molecular Biology - INSACOG | Lamuk Zaveri, Ara Sreenivas, Sofia Banu, Onkar Kulkarni, Shreekant Verma, Amareshwar Vodapalli , Viswagithe S L, B Himasri, Sharath Chandra Thota, Karthik Bharadwaj Tallapaka, Krishna Khairnar, Rakesh K Mishra,Divya Tej Sowpati |
| EPI_ISL_1360361 | CSIR-National Environmental Engineering Research Institute | CSIR-Centre for Cellular and Molecular Biology - INSACOG | Ara Sreenivas, Onkar Kulkarni, Lamuk Zaveri, Sofia Banu, Shreekant Verma, Amareshwar Vodapalli , Viswagithe S L, B Himasri, Sharath Chandra Thota, Karthik Bharadwaj Tallapaka, Krishna Khairnar, Rakesh K Mishra,Divya Tej Sowpati |
| EPI_ISL_1360364 | CSIR-National Environmental Engineering Research Institute | CSIR-Centre for Cellular and Molecular Biology - INSACOG | Sofia Banu, Lamuk Zaveri, Ara Sreenivas, Shreekant Verma, Onkar Kulkarni, Amareshwar Vodapalli ,Viswagithe S L, B Himasri, Sharath Chandra Thota, Karthik Bharadwaj Tallapaka, Krishna Khairnar, Rakesh K Mishra,Divya Tej Sowpati |
| EPI_ISL_1360370 | CSIR-National Environmental Engineering Research Institute | CSIR-Centre for Cellular and Molecular Biology - INSACOG | Ara Sreenivas, Onkar Kulkarni, Lamuk Zaveri, Sofia Banu, Shreekant Verma, Amareshwar Vodapalli , Viswagithe S L, B Himasri, Sharath Chandra Thota, Karthik Bharadwaj Tallapaka, Krishna Khairnar, Rakesh K Mishra,Divya Tej Sowpati |
| EPI_ISL_1552253 | National Virus Reference Laboratory | National Virus Reference Laboratory | Guerrino Macori, Gabriel Gonzalez, Michael Carr, Zoe Yandle, Charlene Bennett, Jonathan Dean, Seamus Fanning, Cillian F De Gascun |
| EPI_ISL_1595896 | KEMPEGOWDA INTERNATIONAL AIRPORT | INSACOG-KA, NIMHANS | Chitra Pattabiraman, Pramada Prasad, Anson Kunjumon George, Ananthapadmanabha Kotambail, Darshan Sreenivas, Chetan G K, Gautham Arunachal Udupi, Anita S Desai, V Ravi |
| EPI_ISL_1595907 | BBMP Urban PHC | INSACOG-KA, NIMHANS | Chitra Pattabiraman, Pramada Prasad, Anson Kunjumon George, Ananthapadmanabha Kotambail, Darshan Sreenivas, Chetan G K, Gautham Arunachal Udupi, Anita S Desai, V Ravi |
| EPI_ISL_1623010 | National Virus Reference Laboratory | National Virus Reference Laboratory | Zoe Yandle, Charlene Bennett, Gabriel Gonzalez, Michael Carr, Jonathan Dean, Cillian F De Gascun |
| EPI_ISL_1647348 | Division of Emerging Infectious Diseases, Bureau of Infectious Diseases Diagnosis Control, Korea Disease Control and Prevention Agency | Division of Emerging Infectious Diseases, Bureau of Infectious Diseases Diagnosis Control, Korea Disease Control and Prevention Agency | Ae Kyung Park, Il-Hwan Kim, Heui Man Kim, Jeong-Min Kim, Jeong-Ah Kim, Chae Young Lee, Jin Sun No, Eun-Jin Kim |
| EPI_ISL_1662307, EPI_ISL_1662308, EPI_ISL_1662316, EPI_ISL_1662318, EPI_ISL_1662322, EPI_ISL_1662328 | Pt. Jawahar Lal Nehru Memorial Medical College, Raipur, Chhattisgarh | Institute of Life Sciences - INSACOG | Sunil K. Raghav, Safal Walia, Arup Ghosh, Atimukta Jha, Amol M. Kanampalliwar, Shifu Aggarwal, Rupesh Dash, Rajeeb Swain, Punit Prasad, INSACOG Consortium, Ajay Parida |
| EPI_ISL_1662410, EPI_ISL_1662420 | BRLSABVM Government Medical College, Rajnandgaon, Chhattisgarh | Institute of Life Sciences - INSACOG | Sunil K. Raghav, Safal Walia, Arup Ghosh, Atimukta Jha, Amol M. Kanampalliwar, Shifu Aggarwal, Rupesh Dash, Rajeeb Swain, Punit Prasad, INSACOG Consortium, Ajay Parida |
| EPI_ISL_1662427, EPI_ISL_1662431, EPI_ISL_1662444 | Capital hospital, Bhubaneswar | Institute of Life Sciences - INSACOG | Sunil K. Raghav, Safal Walia, Arup Ghosh, Atimukta Jha, Amol M. Kanampalliwar, Shifu Aggarwal, Rupesh Dash, Rajeeb Swain, Punit Prasad, INSACOG Consortium, Ajay Parida |
| EPI_ISL_1663250 | BRLSABVM Government Medical College, Rajnandgaon, Chhattisgarh | Institute of Life Sciences - INSACOG | Sunil K. Raghav, Safal Walia, Arup Ghosh, Atimukta Jha, Amol M. Kanampalliwar, Shifu Aggarwal, Rupesh Dash, Rajeeb Swain, Punit Prasad, INSACOG Consortium, Ajay Parida |
| EPI_ISL_1663313, EPI_ISL_1663315, EPI_ISL_1663316, EPI_ISL_1663320 | CIIMS, Bilaspur, Chhattisgarh | Institute of Life Sciences - INSACOG | Sunil K. Raghav, Safal Walia, Arup Ghosh, Atimukta Jha, Amol M. Kanampalliwar, Shifu Aggarwal, Rupesh Dash, Rajeeb Swain, Punit Prasad, INSACOG Consortium, Ajay Parida |
| EPI_ISL_1725188 | Labor Dr. Wisplinghoff - KÃ¶ln | Robert Koch Institute | unknown |
| EPI_ISL_1847409 | Labor Dr. Spranger | Robert Koch Institute | unknown |
| EPI_ISL_2260427, EPI_ISL_2260462, EPI_ISL_2260468, EPI_ISL_2260478, EPI_ISL_2260556, EPI_ISL_2260580 | Bayerisches Landesamt für Gesundheit und Lebensmittelsicherheit (LGL) | Robert Koch Institute | unknown |
| EPI_ISL_2314337 | Laborarztpraxis Dres. med. Walther Weindel & Kollegen | Robert Koch Institute | unknown |

We gratefully acknowledge the following Authors from the Originating laboratories responsible for obtaining the specimens, as well as the Submitting laboratories where the genome data were generated and shared via GISAID, on which this research is based.

All Submitters of data may be contacted directly via www.gisaid.org

Authors are sorted alphabetically.

| Accession ID | Originating Laboratory | Submitting Laboratory | Authors |
|---|---|---|---|
| EPI_ISL_1663380, EPI_ISL_1663409, EPI_ISL_1663418 | NCCS, Pune | Institute of Life Sciences - INSACOG | Sunil K. Raghav, Safal Walia, Arup Ghosh, Atimukta Jha, Amol M. Kanampalliwar, Shifu Aggarwal, Rupesh Dash, Rajeeb Swain, Punit Prasad, INSACOG Consortium, Ajay Parida |
| EPI_ISL_1663495 | Capital hospital, Bhubaneswar | Institute of Life Sciences - INSACOG | Sunil K. Raghav, Safal Walia, Arup Ghosh, Atimukta Jha, Amol M. Kanampalliwar, Shifu Aggarwal, Rupesh Dash, Rajeeb Swain, Punit Prasad, INSACOG Consortium, Ajay Parida |
| EPI_ISL_1663546 | MGM Medical College, Jamshedpur | Institute of Life Sciences - INSACOG | Sunil K. Raghav, Safal Walia, Arup Ghosh, Atimukta Jha, Amol M. Kanampalliwar, Shifu Aggarwal, Rupesh Dash, Rajeeb Swain, Punit Prasad, INSACOG Consortium, Ajay Parida |
| EPI_ISL_1704392, EPI_ISL_1704407, EPI_ISL_1704471, EPI_ISL_1704472, EPI_ISL_1704474, EPI_ISL_1704519, EPI_ISL_1704520, EPI_ISL_1704526, EPI_ISL_1704531, EPI_ISL_1704535, EPI_ISL_1704623, EPI_ISL_1928408, EPI_ISL_1928411, EPI_ISL_1928430, EPI_ISL_1928432, EPI_ISL_1928459, EPI_ISL_1928510 | | | |
| see above | ICMR-National Institute of Virology - INSACOG | NIV Influenza | Dr. Varsha Potdar |
| EPI_ISL_1939891 | NCCS | inStem NCBS | Uma Ramakrishnan Dasaradhi Palakodeti Aswin SaiNarain |
| EPI_ISL_1970498 | ICMR-National Institute of Virology - INSACOG | NIV Influenza | Dr. Varsha Potdar |
| EPI_ISL_2107058 | National Centre For Cell Science - INSACOG | National Centre For Cell Science | Dhiraj Paul, Shivang P. Bhanushali, Mohak P Gujare, Mitali Inamdar, Manoj Kumar Bhat, Ajay Pillai, INSACOG Consortium team, Yogesh Shouche |
| EPI_ISL_2132715 | INSACOG-WB | National Institute of Biomedical Genomics - INSACOG | Arindam Maitra, Bhaswati Bandyopadhyay, Nidhan Kumar Biswas, Tamal Ghosh, Sreedhar Chinnaswamy, Ajay Chakraborti, Saumitra Das |
| EPI_ISL_2157048 | CSIR-Centre for Cellular and Molecular Biology | CSIR-Centre for Cellular and Molecular Biology-INSACOG | Lamuk Zaveri,Tulasi Nagabandi, Ara Sreenivas,Shreekant Verma, Amareshwar Vodapalli ,Blessy B John,Viswagithe S L,B Himasri,Valli Nagalakshmi Undamatla,Payel Mukherjee,Sofia Banu,Archana Bharadwaj Siva,Sharath Chandra Thota,Karthik Bharadwaj Tallapaka,Rakesh K Mishra,Divya Tej Sowpati |
| EPI_ISL_2231661 | AP SSO | CSIR-Centre for Cellular and Molecular Biology-INSACOG | Tulasi Nagabandi, Lamuk Zaveri,Ara Sreenivas,Shreekant Verma, Amareshwar Vodapalli ,Blessy B John,Viswagithe S L,B Himasri,Valli Nagalakshmi Undamatla,Payel Mukherjee,Sofia Banu,Archana Bharadwaj Siva,Sharath Chandra Thota,Karthik Bharadwaj Tallapaka,Rakesh K Mishra,Divya Tej Sowpati |
| EPI_ISL_2342916, EPI_ISL_2342925, EPI_ISL_2342933, EPI_ISL_2342949, EPI_ISL_2342962 | NCCS, Pune | Institute of Life Sciences - INSACOG | Sunil K. Raghav, Safal Walia, Arup Ghosh, Atimukta Jha, Amol M. Kanampalliwar, Omprakash Shiriwas, Sana Fatma, Shifu Aggarwal, Rupesh Dash, Rajeeb Swain, Punit Prasad, INSACOG Consortium, Ajay Parida |
| EPI_ISL_2373285 | CSIR-Centre for Cellular and Molecular Biology | CSIR-Centre for Cellular and Molecular Biology-INSACOG | Blessy B John,Viswagithe S L,Tulasi Nagabandi, Lamuk Zaveri,Ara Sreenivas,Shreekant Verma, Amareshwar Vodapalli ,B Himasri,Valli Nagalakshmi Undamatla,Payel Mukherjee,Sofia Banu,Archana Bharadwaj Siva,Sharath Chandra Thota,Karthik Bharadwaj Tallapaka,Rakesh K Mishra,Divya Tej Sowpati |
